# Effects of individual components of remote monitoring for implantable cardioverter defibrillators or cardiac resynchronisation therapy defibrillators: protocol for a systematic review and component network meta-analysis

**DOI:** 10.1101/2024.03.07.24303950

**Authors:** Makiko Okazaki, Natsuko Sekiguchi, Yuki Sahashi, Hisashi Noma, Takahiro Mihara

**Affiliations:** Department of Health Data Science, Yokohama City University Graduate School of Data Science, Yokohama, Japan; Department of Clinical Engineering, Sakakibara Heart Institute, Fuchu, Japan; Division of Nursing, Higashigaoka Faculty of Nursing, Tokyo Healthcare University, Meguro, Japan; Department of Cardiology, Cedars-Sinai Medical Center, Los Angeles, USA; Department of Cardiology, Gifu University, Gifu, Japan; Department of Data Science, The Institute of Statistical Mathematics, Tachikawa, Japan

**Keywords:** Remote Monitoring, Defibrillators, Implantable, Cardiac Resynchronisation Therapy Devices, Network Meta-Analysis, Outpatients, Hospitalization, Randomized Controlled Trials

## Abstract

**Introduction:** Currently, the standard of care for patients with cardiac implantable electronic devices (CIEDs) such as implantable cardioverter-defibrillators (ICD) and cardiac resynchronisation therapy-defibrillators (CRT-D) involves a combination of in-person outpatient visits and remote monitoring (RM). RM consists of scheduled remote device interrogation and automated transmission of prespecified alerts (alert transmission) at varying frequencies depending on manufacturers and institutions.

However, the effects of RM factors on prognosis remain unclear. This systematic review and component network meta-analysis (CNMA) will aim to investigate which RM components (device interrogation, alert transmission, and data transmission frequency) have the greatest impact on prognosis in patients with ICD or CRT-D.

**Methods and analysis:** A systematic review will be conducted using MEDLINE (PubMed), Embase, the Cochrane Central Register of Controlled Trials, the Web of Science, Clinical Trials.gov, the WHO International Clinical Trials Registry Platform (ICTRP), the European Union Clinical Trials Register (EU-CTR), and the University Hospital Medical Information Network Clinical Trials Registry (UMIN-CTR). We will include randomised controlled trials (RCTs) assessing the effect of RM on patient outcomes in individuals with ICD or CRT-D. The primary outcome will be hospitalisation due to cardiovascular disease, heart failure, and device-related complications. Three reviewers will independently screen the titles and abstracts of identified studies. Two reviewers will extract data independently, and risk of bias will be assessed by one reviewer and verified by a second. Random-effects model pairwise meta-analysis, random-effects network meta-analysis (NMA), and additive CNMA will be applied in data synthesis. To assess the quality of evidence, we will employ the Grading of Recommendations Assessment, Development, and Evaluation (GRADE) approach for pairwise meta-analysis and the Confidence in Network Meta-Analysis (CINeMA) approach for NMA.

**Ethics and dissemination:** Ethics approval is not required as this study will use existing published data. The results will be submitted for publication in a peer-reviewed journal.

**PROSPERO registration number**: CRD42024517406.

## INTRODUCTION

### Rationale

Implantable cardioverter-defibrillator (ICD) and cardiac resynchronisation therapy-defibrillator (CRT-D) implantation improves cardiac function and prevents sudden arrhythmic death [1, 2]. The current post-implantation standard of care involves a combination of in-person outpatient visits and remote monitoring (RM) [3, 4]. To date, several studies have examined the safety and efficacy of RM [4–6] and reported that RM reduces the number of in-person outpatient visits without compromising safety [5], decreases all-cause mortality [6], and reduces emergency clinic visits in patients with heart failure and ICD or CRT-D implants [7]. These studies have predominantly compared RM with conventional in-person outpatient care.

RM involves scheduled remote device interrogation and automated transmission of prespecified alerts (alert transmission) [8]. Furthermore, the frequency of remote data transmission varies depending on the manufacturers and institutions involved in reported studies [9]. The frequency of in-person outpatient visits also differs between patients with and without RM, with RM often allowing for extended intervals [5, 7, 8]. In addition, the combination of in-person outpatient visits, scheduled remote device interrogation, and alert transmission demands substantial staff time [10]. Therefore, identifying which RM components (i.e., scheduled interrogation and alert transmission, including interrogation intervals) have the greatest impact on patient outcomes is crucial to understanding which component should be prioritised in patient management. However, the effects of RM components on outcomes and patient management remain unclear.

Component network meta-analysis (CNMA) allows the estimation of the individual effects of multiple components within complex interventions [11]. Therefore, the study will utilise CNMA to elucidate the relative effects of remote device interrogation, alert transmission, and data transmission frequency on patient outcomes.

### Objectives

The objective of this systematic review will be to evaluate which components of RM have the greatest impact on prognosis in patients with ICD or CRT-D.

## METHODS

### Study design

The study will be a systematic review incorporating CNMA. This protocol follows the Preferred Reporting Items for Systematic Review and Meta-Analysis Protocols (PRISMA-P) guidelines [12]. The systematic review will be reported using the PRISMA guidelines extension for systematic reviews incorporating network meta-analyses of health care interventions (PRISMA-NMA) [13] to structure the contents of the final report. We will conduct CNMA according to PRISMA-NMA guidelines and the *Cochrane Handbook for Systematic Reviews of Interventions* version 6.4 [13, 14]. This protocol has been registered with the International Prospective Register of Systematic Reviews (PROSPERO) (registration number: CRD42024517406).

### Eligibility criteria

We will include randomised controlled trials (RCTs), including re-analyses of previously published RCTs that did not originally include the outcomes covered by this study. We will exclude quasi-experimental studies.

### Participants

We will include studies examining patients who underwent ICD or CRT-D implantation and studies involving other cardiac implantable electronic devices (CIEDs) if ICD or CRT-D data are reported separately.

### Interventions

RM involving data transmission without in-person interaction will be included. RM will comprise scheduled interrogation and alert transmission. In clinical practice, the schedule of remote interrogations is typically determined by treating physicians in accordance with clinical guidelines [3,4]. Thus, in CNMA, intervention components will be classified as short-interval remote interrogation (<1 month), long-interval remote interrogation (≥1 month), or alert transmission.

### Comparators

Conventional in-person outpatient visits will be the reference component, as these are the most common comparators reported in published studies and are routinely performed in clinical practice. The frequency of in-person outpatient visits differs between patients with and without RM, with RM often allowing longer intervals.

Therefore, in CNMA, in-person outpatient visits will be classified as short-interval in-person outpatient visits (<12 months) and long-interval in-person outpatient visits (≥12 months).

### Outcomes

Studies including at least one of the following outcomes are eligible including outcomes reported as a component of a composite outcome:

- hospitalisation (cardiovascular, heart failure, device-related)
- mortality (all-cause, cardiovascular)
- unscheduled outpatient visits
- unscheduled hospitalisation

### Timing

No restrictions on follow-up periods will be applied.

### Setting

No restrictions on setting will be applied.

### Language

We will not impose language restrictions in our literature searches.

### Information sources

We will use MEDLINE (PubMed), Embase, the Cochrane Central Register of Controlled Trials, and the Web of Science. The reference lists of the relevant articles will also be searched. Further, we will conduct searches via Clinical Trials.gov, the WHO International Clinical Trials Registry Platform (ICTRP), the European Union Clinical Trials Register (EU-CTR), and the University Hospital Medical Information Network Clinical Trials Registry (UMIN-CTR).

### Search strategy

Search strategies will be developed using medical subject headings (MeSH) and free text words relating to ICD, CRT-D, and RM. The proposed search strategy for MEDLINE is presented in Table 1. Search strategies for other databases, registries, and websites are explained in Supplementary File 1.

**Table 1.** Search strategy for PubMed

| Number | Search terms |
| --- | --- |
| #1 | "defibrillators, implantable"[MeSH Terms] OR<br>("implantable"[Title/Abstract] AND "cardioverter*"[Title/Abstract]) OR<br>("implantable"[Title/Abstract] AND "defibrillator*"[Title/Abstract]) OR<br>("implantable"[Title/Abstract] AND "cardioverter"[Title/Abstract] AND<br>"defibrillator*"[Title/Abstract]) OR "cardiac resynchronization therapy<br>devices"[MeSH Terms] OR ("cardiac"[Title/Abstract] AND<br>"resynchronization"[Title/Abstract]) OR ("cardiac"[Title/Abstract] AND<br>"resynchronisation"[Title/Abstract]) OR<br>("resynchronization"[Title/Abstract] AND "therapy"[Title/Abstract]) OR<br>("resynchronisation"[Title/Abstract] AND "therapy"[Title/Abstract]) OR<br>("resynchronization"[Title/Abstract] AND "device*"[Title/Abstract]) OR<br>("resynchronisation"[Title/Abstract] AND "device*"[Title/Abstract]) OR |
|  | "biventricular"[Title/Abstract] OR "CIED*"[Title/Abstract] OR<br>("biventricular"[Title/Abstract] AND "implantable"[Title/Abstract] AND<br>"cardioverter"[Title/Abstract] AND "defibrillator*"[Title/Abstract]) OR<br>("cardiac"[Title/Abstract] AND "implantable"[Title/Abstract] AND<br>"electronic"[Title/Abstract] AND "device*"[Title/Abstract]) |
| #2 | ("remote"[Title/Abstract] AND "monitor*"[Title/Abstract]) OR<br>("remote"[Title/Abstract] AND "manag*"[Title/Abstract]) OR<br>("remote"[Title/Abstract] AND "follow up"[Title/Abstract]) OR<br>("remote"[Title/Abstract] AND "transmission"[Title/Abstract]) OR<br>("wireless"[Title/Abstract] AND "monitor*"[Title/Abstract]) OR<br>("wireless"[Title/Abstract] AND "manag*"[Title/Abstract]) OR<br>("wireless"[Title/Abstract] AND "follow up"[Title/Abstract]) OR<br>("wireless"[Title/Abstract] AND "transmission"[Title/Abstract]) OR<br>"homemonitor*"[Title/Abstract] OR<br>"remotemanagement"[Title/Abstract] OR "home<br>monitor*"[Title/Abstract] OR "telemonitor*"[Title/Abstract] OR<br>"Telemetry"[MeSH Terms] OR "Telemetry"[Title/Abstract] OR<br>"telemedicine"[MeSH Terms] OR "telemedicine"[Title/Abstract] |
| #3 | ("randomized controlled trial"[Publication Type] OR "controlled clinical<br>trial"[Publication Type] OR "randomized"[Title/Abstract] OR<br>"placebo"[Title/Abstract] OR "drug therapy"[MeSH Subheading] OR<br>"randomly"[Title/Abstract] OR "trial"[Title/Abstract] OR<br>"groups"[Title/Abstract]) NOT ("animals"[MeSH Terms] NOT |
|  | "humans"[MeSH Terms]) |
| #4 | #1 AND #2 AND #3 |

### Study records

#### Data management

Database citations will be exported using Mendeley Reference Manager (https://www.mendeley.com/reference-management/reference-manager; Elsevier, Amsterdam, Netherlands). The search results will be uploaded to Rayyan (http://rayyan.qcri.org; Rayyan Systems, Cambridge, MA, USA), a free web and mobile application that facilitates abstract and title screening and collaboration among reviewers [15].

### Study selection

Three reviewers will independently screen the titles and abstracts of each study identified using the described search strategies. We will retrieve the full texts of studies that appear to meet the eligibility criteria and those of which eligibility is questioned. Disagreements between reviewers will be resolved through discussion.

### Data collection process

Two reviewers will independently extract data in duplicate from each eligible study. To ensure consistency between reviewers, the first 5 titles will be screened via a data collection form and discussion. Discrepancies will be resolved via discussion (including a third reviewer if necessary). If data are missing or presented ambiguously, we will contact the study authors for clarification.

### Data items

A data collection sheet will be created. The following data will be extracted:

1. patient characteristics (age, sex, device type, New York Heart Association functional class, underlying heart disease, and left ventricular ejection fraction)
2. intervention and control details (RM interrogation schedule, in-person outpatient visit timing, and alert transmission criteria)
3. outcome data (including mortality, hospitalisation, and dropout rate)
4. study details (title, author information, year of publication, trial design, trial size, eligibility criteria, exclusion criteria, duration of follow-up, type and source of financial support, study settings, and publication status)

Values, for example, means, will be approximated from figures if necessary.

### Outcomes and prioritisation

#### Primary outcome

Hospitalisation including cardiovascular, heart failure, and device-related hospitalisation.

#### Secondary outcomes

All-cause mortality, cardiovascular mortality, unscheduled outpatient visits, and unscheduled hospitalisation.

If outcomes are reported as a composite endpoint, we will extract individual outcomes of interest from study results if extractable. If not extractable, we will contact the study authors for clarification.

#### Risk of bias in individual studies

One reviewer will assess the risk of bias, and a second reviewer will verify the assessment to confirm agreement. Any disagreements will be resolved through discussion. The revised Cochrane risk-of-bias tool for RCTs (RoB 2.0; cochrane.org) [16] will be used. The tool assesses bias across 5 domains, and the overall risk of bias is determined based on the results of these domains:

1. Bias arising from the randomisation process
2. Bias due to deviations from intended interventions
3. Bias due to missing outcome data
4. Bias in the measurement of outcomes
5. Bias for selection of the reported result

Each domain’s risk of bias and overall risk of bias will be described as “low,” “some concern,” or “high.”

### Data synthesis and analysis

#### Summary measures (measures of treatment effect)

Dichotomous outcomes, such as hospitalisation and mortality, will be analysed using risk ratio (RR) with 95% confidence intervals (CI). Continuous outcomes will be analysed using mean difference (MD) or standardised mean difference (SMD) if different measurement scales are used, with 95% CI. If numerical outcome data are not reported in the main text, values will be extracted from figures or tables. If data is missing, we will contact the authors of the study to obtain the relevant missing data.

For unscheduled outpatient visits, several effect measures may be available (risk ratio, mean number of visits per patient, and incidence rate ratio). However, because substantial variation in follow up duration across studies is anticipated, we consider incidence rate ratios (IRRs) to be the most appropriate summary measure and will therefore use them for pooling. When outcomes are reported as incidence rates accompanied only by p-values, standard errors will be derived by applying distributional assumptions appropriate to the statistical test used.

#### Pairwise meta-analysis

If multiple studies with conventional in-person outpatient visits as a comparator are identified, we will conduct pairwise meta-analyses to assess the effectiveness of RM intervention for each outcome. We will use the Hartung-Knapp-Sidik-Jonkman [17,18,19] meta-analysis with random effects method to combine the results and present summary measures alongside the estimated effects of each study using forest plots. Heterogeneity will be quantified using *I²* statistics.

### Network meta-analysis

#### Review of network geometry

We will construct a network diagram and evaluate the network geometry [13]. Evaluating the geometry of the network allows for an assessment of the feasibility of NMA, such as determining whether the network of interventions is connected.

Additionally, this assessment includes the identification of closed loops of treatments within the network, facilitating the evaluation of inconsistency that is the disagreement between effects estimated from direct and indirect sources.

#### Transitivity and inconsistency in NMA

We will statistically evaluate both local and global inconsistency. The local assessment will be performed using the side-splitting method [20] while the global assessment will be conducted via the design-by-treatment interaction model [21].

We will perform a random-effects NMA assuming a common between-studies variance across the whole network. Summary effect measures such as RR will be estimated along with 95% CI. The results of the estimation will be presented using the league table and the Surface Under the Cumulative Ranking curve (SUCRA) [22], or the P-score, a frequentist version of SUCRA[23]. We will use the R package “nma” (cran.r-project.org/web/packages/NMA/index.html) to estimate SUCRA [24], and the R package “netmeta” (cran.r-project.org/web/packages/netmeta/index.html) to calculate P-score [25].

### Component network meta-analysis

#### Additivity assumption in CNMA

CNMA allows the estimation of component effects of multicomponent interventions. In this context, an additivity assumption is used, which means that the effect of each intervention can be expressed as the sum of the effects of its individual components. We will use the method based on a comparison of treatment estimates from the standard NMA and the additive CNMA model to assess the additivity assumption [11,26].

If additivity holds, we will use an additive effects-based CNMA model for estimating the relative effects of components. The results of the estimation will be presented using the league table and the P-score [23]. We will use the R package “netmeta” (cran.r-project.org/web/packages/netmeta/index.html) [25].

### Narrative synthesis

If quantitative synthesis is not feasible due to significant between-studies heterogeneity or an insufficient number of studies, we will conduct systematic narrative synthesis.

This approach will use information from the text and tables to summarise and describe the characteristics and findings of the included studies.

### Additional analyses

#### Sensitivity analysis

To assess the robustness of our findings based on the primary analysis, we plan to perform a sensitivity analysis after excluding studies with a high risk of bias.

#### Subgroup analysis

We will conduct a subgroup analysis classified by device type (ICD or CRT-D) to examine the consistency of results and validate the robustness of our findings.

Statistical analyses will be performed using the latest versions of R software (R Foundation for Statistical Computing, Vienna, Austria) [27] and RStudio (RStudio, Boston, MA, USA) [28] at the time of analysis.

### Risk of bias across studies

If a study protocol is available, we will compare outcomes reported in the protocol or trial registry with those in the published studies to assess the potential risk of reporting bias. Small study effects will be assessed using Egger’s regression to detect funnel plot asymmetry [29] using a significance threshold of *p*<0.1.

### Confidence in cumulative estimate

We will use the Grading of Recommendations Assessment, Development, and Evaluation (GRADE) approach to assess the certainty of evidence for each outcome in pairwise meta-analyses [30]. The quality of evidence will be assessed across the following domains: limitations in study design, risk of bias, inconsistency, indirectness, imprecision of the results, and publication bias. The quality of evidence will be categorised as high, moderate, low, or very low. The Confidence in Network Meta-Analysis (CINeMA) approach will be used to evaluate confidence in the NMA estimates [31–33].

## IMPLICATION AND LIMITATION

In the proposed study, we will aim to estimate the effect size of each RM component on patient outcomes. The current follow-up practices for CIEDs, including ICDs and CRT-Ds, recommend RM [3,4] based on known benefits including reductions in all-cause mortality and emergency clinic visits [6, 7]. However, managing the increasing population of patients with CIEDs using conventional remote management (periodic remote interrogation + alert transmission + in-person outpatient visits) is a major clinical and administrative burden [34]. Therefore, by identifying the most effective components of RM, follow-up can be optimised without compromising patient outcomes. However, CNMA assumes both consistency and additivity. If these assumptions are not met, the accuracy of the estimated results will potentially be compromised, therefore necessitating careful interpretation.

## ETHICS AND DISSEMINATION

Ethics approval is not required as this study will use existing published data. The results will be submitted for publication in a peer-reviewed journal.

Any significant changes to this protocol will be noted with a description of the change, the corresponding rationale, and the date of the amendment when the results are reported.

## Data Availability

Data sharing not applicable as no datasets were generated and/or analysed for this study.

## Acknowledgments

We would like to thank Editage (www.editage.com) for English-language editing.

## Data availability statement

Data sharing not applicable as no datasets were generated and/or analysed for this study.

## Authors’ contributions

TM drafted the protocol. MO, TM, and YS led the development of the review protocol and drafted the manuscript. TM and MO contributed to the development of the selection criteria, risk of bias assessment strategy and data extraction criteria. TM and MO developed the search strategy. HN provided expertise on statistical analysis. MO, NS, YS, and TM read all drafts of the manuscript, provided feedback and approved the final manuscript.

## Registration

In accordance with the guidelines, our systematic review protocol was registered with the International Prospective Register of Systematic Reviews (PROSPERO) on 7 March 2024 (registration number CRD42024517406).

